# Mendelian Randomization Identifies Circulating Proteins APOM and TNXB as Biomarkers for Steroid Sensitive Nephrotic Syndrome

**DOI:** 10.1101/2025.10.03.25337251

**Authors:** Daniel Heydari, Sheldon Langlois, Mahboobeh Norouzi, Robert L Myette, Susan Samuel, Sirui Zhou, Tomoko Takano, Guillaume Butler-Laporte, Mallory L Downie

## Abstract

**Introduction:** Steroid-sensitive nephrotic syndrome (SSNS) is the most common glomerular disease in children worldwide. Current treatments are not targeted and lead to serious adverse effects. We sought to identify circulating proteins associated with SSNS in European children using an unbiased two-sample Mendelian randomization (MR) and colocalization approach to inform novel drug targets for disease.

**Methods:** We conducted a large-scale MR study using *cis* genetic determinants (protein quantitative trait loci, pQTL) of 1,540 circulating proteins from eight large genome-wide association studies to screen for causal association of these proteins with SSNS risk in 422 children with SSNS and 5642 control subjects. We then performed genetic colocalization to further investigate loci identified by MR.

**Results:** We found four proteins causally linked to SSNS by MR, two of which colocalized. We found that genetically predicted increases in apolipoprotein M (APOM) level and Tenascin XB (TNXB) level were associated with decreased risk of SSNS [p = 1.37×10^−5^; MR Odds Ratio (OR) 0.40, 95% CI 0.27-0.61 for APOM, and p = 2.95×10^−4^; OR 0.49, 95% CI 0.33-0.72 for TNXB]. Colocalization with SSNS occurred at *HLA-DRB1* (98%) and *HLA-DQA1* (79%) for APOM and TNXB, respectively. Follow-up binding affinity and gene expression analysis showed that APOM and TNXB peptides have high binding affinity for their respective HLA-pQTLs, and that APOM has a biologically plausible causal relationship with SSNS.

**Conclusions:** We identified two novel blood proteins associated with steroid sensitive nephrotic syndrome in children using an MR and colocalization approach. These biomarkers are promising targets for development of drugs and/or screening tools for early prediction of disease.

## INTRODUCTION

Steroid sensitive nephrotic syndrome (SSNS) is the most common glomerular disease in children worldwide, with incidence of up to 16 per 100,000 children.^1^ It is characterized by massive proteinuria resulting from injury to podocytes, the specialized cell which forms the glomerular filtration barrier.^1^ Children with SSNS are at risk for thrombosis, sepsis, and progression to end stage kidney disease.^1,2^ Corticosteroids are the mainstay of treatment but are associated with severe adverse effects.^3^ In 80% of children, SSNS relapses, requiring repeated treatments with corticosteroids.^1^ Much of the morbidity associated with SSNS is related to the toxicity of treatment with corticosteroids, which are non-specific in action.^4^ Thus, novel validated targets are needed for the development of targeted, effective, less toxic treatments in SSNS.

Circulating proteins are one such source of promising new targets. Recent large-scale proteomics research has enabled the measurement of thousands of circulating proteins in the blood. Combining these data with human genetics greatly improves the probability of successful development of new drugs.^5^ Furthermore, these proteomics data are widely available, and using them to identify drug targets in disease can accelerate the translation of this data to new therapies for patients.^6^

Recent discovery of anti-nephrin antibodies and antibodies against other slit diaphragm components in a subset of children with SSNS have transformed our understanding of disease pathogenesis,^7–9^ but the precise immune derangements that predispose to antibody production or explain disease onset in antibody-negative children remain unclear. Furthermore, determining the causal factors of SSNS via observational case-control studies of antibody and/or cytokine profiling is challenging due to the presence of confounding and reverse causation factors.

Mendelian randomization (MR) is a genetic epidemiology method that uses genetic variation to answer a causal question about how a modifiable exposure (here, circulating proteins) affects an outcome (here, SSNS).^10^ Genetic variants are randomly allocated at conception, and thus, minimally influenced by environmental confounding factors.^11^ Similarly, a disease state does not typically alter genetic variation.^12^ Thus, MR avoids the inherent limitations of traditional observational studies, such as confounding and reverse causation, both of which compromise causal inference in observational study design.^13^

Two-sample MR tests for causal associations using genetic variants obtained from genome-wide association studies (GWAS) in two separate populations.^14^ This method often improves statistical power compared to one-sample because it allows for larger sample sizes for exposure and outcome GWAS.^15^ Causal associations are then detected if the exposure and outcome have shared causal variants (single nucleotide variants, SNVs) at the same locus. However, a causal association may be confounded by linkage disequilibrium (LD),^12^ thus genetic colocalization must also be performed on MR-prioritized signals.

In this study, we used two-sample MR to screen the genetic variants associated with over 1500 previously measured blood proteins altering the risk of SSNS. We identified *cis-*acting SNVs for blood proteins (*cis*-protein quantitative trait loci [*cis*-pQTL], meaning SNVs adjacent to or within the genes that encode the proteins) in eight large proteomic GWAS. We then used these *cis*-pQTL to estimate the effect of genetically altered levels of their corresponding proteins on risk of SSNS in Europeans. We then performed colocalization using HLA alleles, reverse MR, and downstream analysis of priority targets.

## METHODS

The methods used to conduct this MR study in SSNS adhered to the STROBE guidelines.^14^

### Study Exposure: Proteomic GWAS

We identified *cis-*pQTLs from eight large proteomic GWAS using European populations only.^16–22^ Niu et al was performed in children <18 years, whereas the other seven GWAS included adults >18 years. Circulating proteins from Sun et al (2018),^23^ Ferkingstad et al,^17^ Pietzner et al,^19^ Suhre et al,^24^ and Emilsson et al^18^ were measured on the SomaLogic platform; Sun et al (2023) and Yao et al,^16,21^ used protein measurements on the O-link platform. Niu et al used a mass spectrometry approach to quantify proteins in plasma samples.^22^ See Supplementary Data 1 for information on the number of study participants, proteins measured, and *cis*-pQTLs identified in each study.

### Study Outcomes: SSNS GWAS

To assess the association of *cis*-pQTLs with SSNS outcomes, we used the largest available SSNS GWAS in Europeans, which comprised 422 pediatric patients and 5,642 healthy controls.^25^

### Two-sample MR

We performed two-sample MR using the TwoSampleMR R package v0.6.20 to screen potential circulating blood proteins for their role in influencing SSNS susceptibility.^10^ We used the Wald ratio to estimate the effect of each protein on SSNS. We selected the lead SNVs (*cis*-pQTLs) for association with protein levels in the eight proteomic GWAS (separating child and adult studies). We then queried the lead *cis*-pQTL for association with SSNS in the GWAS by Dufek et al.^25^ Bonferroni correction was used to control for multiple comparisons between the total number of distinct proteins tested (*p* = 3.4×10^−5^ = 0.05/1,465 in adults, and *p* = 2.9 × 10^−4^ = 0.05/171 in children). The results are presented as MR odds ratio [OR] with 95% confidence intervals for risk of SSNS per genetically predicted 1 standard deviation change in circulating protein level.

### MR Assumptions

MR studies rely on three core assumptions.^26^ First, MR requires a robust association between genetic variants and the circulating protein of interest.^26^ This is established by choosing only those variants (*cis*-pQTLs) achieving genome-wide significant (*p* < 5×10^−8^) association with the protein, which we have done in this study. We also included independent *cis-*pQTLs, selected via LD clumping. We used the *ld_clump* function from ieugwasr v1.1.0^27^ and the 1000 Genomes European population reference panel^28^ to clump and select LD-independent *cis*-pQTLs (R^2^ < 0.01, with a clumping window of 250kb). We included the same proteins represented by different *cis*-pQTLs from difference studies to cross-examine our findings. For *cis*-pQTLs that were not present in the SSNS GWAS, we queried for proxy SNVs that were in high LD (R^2^ > 0.8, MAF < 0.42) using the *LDproxy* function from LDlinkR v1.4.0.^29^ *cis*-pQTLs with ambiguous effects were removed before MR to prevent allele mismatches and false positive findings.^29^ *cis*-pQTLs for multiple proteins and rare *cis*-pQTLs (MAF < 0.01) were also removed. As an additional metric of strength, we only included *cis*-pQTLs with an *F-*statistic > 10, which is an indication of a strong MR instrument.^26^ F-statistic was calculated using the formula F = [r^2^/k] / [(1-r^2^)/(n - k −1)], where r^2^ is the proportion of variance of the protein level explained by its *cis*-pQTL, k is the number of instruments used in the model, and n is the GWAS sample size.^30^

The second MR assumption is that the genetic variants are not associated with other confounding factors – the most important being population stratification (genetic differences due to ancestry).^26^ All case and control subjects in the proteomic GWAS and in the SSNS GWAS were of European ancestry, determined by genetic principal component analysis, which minimizes the likelihood of confounding due to population structure.

The third MR assumption is horizontal pleiotropy, which occurs when the genetic variant itself directly affects the outcome, rather than through the exposure.^26^ Potential bias due to horizontal pleiotropy is greatly reduced in our study since we used *cis*-acting SNVs, meaning within 1Mb of the affected protein’s promoter region.^14^ *Cis*-pQTLs are unlikely to impact the disease independently of the protein levels because they are physically close to the gene that encodes the protein.^16^ To further eliminate bias by horizontal pleiotropy, we used PheWeb (https://pheweb.org/) to identify any *cis*-QTLs that were previously associated with any potential confounders (*p* < 5 × 10^−8^).^31,32^ For *cis*-pQTLs of proteins prioritized by MR and measured on the SomaLogic platform, we also queried the potential for aptamer-binding effects. Aptamer-binding effects occur when the *cis-*pQTL itself or a nearby protein-altering variant (PAV) affect protein-protein interactions, and as a consequence, affects the accuracy of protein measurements.^19^ This is important because PAVs can influence protein binding to antibodies used in ELISA assays, which would be needed for future validation studies of a candidate protein.^19^

### Reverse Causality Testing

For each MR-prioritized protein, we also prioritized those for which Steiger directionality was TRUE. Steiger filtering tests if a *cis*-pQTL explains more of the variance in the exposure than the outcome.^15^ Retaining instruments with a greater effect on the exposure reduces the likelihood of a false positive result and of violating MR assumptions due to reverse causality.^15^

Reverse MR was conducted for MR-prioritized proteins to further investigate reverse causality. The same methods were used as documented above, except that the SSNS GWAS was used as the exposure and the MR-prioritized protein GWAS were used as the outcome. Reverse MR were multi-instrument analyses, and thus an inverse-variance weighted approach as well as various pleiotropy-robust MR methods (MR-Egger, weighted median, weighted mode, and MR PRESSO) were used.

### eQTL Assessment

We tested for evidence of being an expression quantitative trait loci for the *cis*-pQTLs of all MR-prioritized proteins using GTex.^33^ We also queried tissues expression of MR-prioritized in kidney cells using the Kidney Cell Atlas.^34^

### Colocalization

To test for confounding by LD, all loci harboring the *cis*-pQTLs of the MR-prioritized proteins were tested for colocalization with SSNS, using coloc v6.0.0 and HLAcoloc v2.1.^35–37^ Genetic colocalization via SNVs is the standard method used to evaluate confounding by LD, however, at the HLA locus (chromosome 6:28,510,120-33,480,577), using colocalization via SNVs is often computationally impossible due to the high degree of LD at the HLA.^37^ In our study, pQTLs for MR-prioritized proteins were located within the HLA locus, and thus, in addition to coloc, we employed a novel colocalization method using HLA alleles to colocalize, rather than SNVs.^37^ Similar to SNV-based colocalization which uses Bayesian analysis to calculate posterior inclusion probabilities, colocalization via an HLA allele with posterior probability for hypothesis 4 (PP4, that there is association for both protein level and SSNS outcome, and that they are driven by the same causal HLA allele) > 0.5 were considered likely to colocalize, and PP4 > 0.8 were considered highly likely to colocalize.^37^

For SNV colocalization, we analyzed SNVs with minor allele frequency of >0.01 within 1Mb (± 500kb) of each *cis*-pQTL of interest. An LD matrix of the corresponding SNVs was generated by the *ld_matrix* function from ieugwasr v1.1.0 (1000 Genomes Project European reference panel).^27,28^ For HLA colocalization, we analyzed HLA alleles from the HLA-pQTL summary level data, recently published by Krishna et al.^38^ SNVs and HLA alleles for SSNS were obtained directly from the Dufek et al GWAS,^25^ and an LD matrix of the HLA alleles was generated using ieugwasr and an HLA allele reference panel from the Type 1 Diabetes Genetics Consortium. ^25,27,39^

### HLA binding assessment

For each HLA allele identified by colocalization for MR-prioritized proteins, we evaluated binding affinity of peptides within these proteins using the NetMHCIIpan resource.^40^ NetMHCIIpan is a software tool that predicts the binding affinity of peptides to major histocompatibility complex (MHC) molecules.^40^ We used this tool to evaluate whether the HLA alleles identified as pQTLs for our MR-prioritized proteins are likely to present peptides derived from these proteins. HLA molecules can affect blood protein levels by altering the natural immune response or affecting protein expression in immune cells.^37^ Predicting strong binding affinity to MHC molecules with peptides of our MR-prioritized proteins would further add support to both the tested HLA-pQTLs and to our HLA-colocalization results.

Since both MR-prioritized proteins colocalized with genetic signals at HLA class II loci, we used NetMHCIIpan to predict the binding affinities of peptides derived from these proteins to all available HLA class II molecules. We compared predicted affinities across alleles, expecting that the allele identified as the colocalized HLA-pQTL would show higher predicted binding affinity to peptides from the corresponding protein. We tested for multiple peptide lengths (10, 15, and 20) and set the threshold for strong binders and weak binders as the top 1% and 5%, respectively.

### Power analysis

When we have strong SNV-instruments from the exposure GWAS, statistical power in an MR study becomes a function of the sample size of the outcome (SSNS) GWAS, as well as the variance of the circulating protein levels determined by the protein-increasing allele (or risk allele) of the variant.^41^ We calculated power for our study using the tool published by Brion et al.^42^

### Data Availability

All data used in this study were obtained from previously published studies. Proteomics datasets are available from the referenced peer-reviewed publications. HLA-pQTL datasets are available from corresponding authors on request. Summary statistics from the SSNS GWAS are publicly available from the GWAS Catalog. The code used to produce these results is available at github.com/orgs/Downie-Lab/.

## RESULTS

### MR Study

The study design is illustrated in Figure 1. We obtained genome-wide significant *cis-*pQTLs or highly correlated proxies (R^2^>0.8) from eight large proteomic GWAS of European ancestry: 1003 proteins from Sun et al (2023),^16^ 697 proteins from Ferkingstad et al,^17^ 82 proteins from Emilsson et al,^18^ 434 proteins from Pietzner et al,^19^ 25 proteins from Sun et al (2018),^23^ 12 proteins from Suhre et al,^24^ 25 proteins from Yao et al,^21^ and 218 proteins from Niu et al.^22^ A total of 1,672 *cis*-pQTLs and 86 LD proxies in adults, and 218 *cis*-pQTLS and 10 LD proxies in children were used as genetic instruments for circulating proteins in SSNS (n=422 cases and 5642 controls).^25^ Since generally there was a single *cis-*pQTL per protein, we performed single-instrument two-sample MR using the Wald ratio to estimate the effect of each candidate protein on SSNS susceptibility. MR analyses revealed that levels of three circulating proteins - human leukocyte antigen E (HLA-E), complement component 4A (C4A), and apolipoprotein M (APOM) - were found to be associated with SSNS using the adult-derived proteomic data (p = 2.95×10^−7^; OR 8.19, 95% CI 3.67 – 18.3 for HLA-E, p = 1.63×10^−6^; OR 0.12, 95% CI 0.05–0.61 for C4A, and p = 1.37×10^−5^; OR 0.40, 95% CI 0.27–0.61 for APOM), and two proteins - complement component 4A (C4A) and Tenascin XB (TNXB) - were associated with SSNS using the pediatric-derived proteomic data (p = 9.82×10^−10^; OR 0.44, 95% CI 0.34–0.57 for C4A and p = 2.95×10^−4^; OR 0.49, 95% CI 0.33– 0.72 for TNXB) after Bonferroni false discovery rate correction (Tables 1-2, Figure 2, and Supplementary Data 2-7). Higher blood levels of C4A, TNXB, and APOM conferred a protective effect (reduced likelihood of SSNS development), whereas higher HLA-E levels increased SSNS risk. All *cis*-pQTLs for these candidate proteins had an F statistic >10, implying that they were strong MR instruments (Supplementary Data 2 and 5).

**Figure 1.**
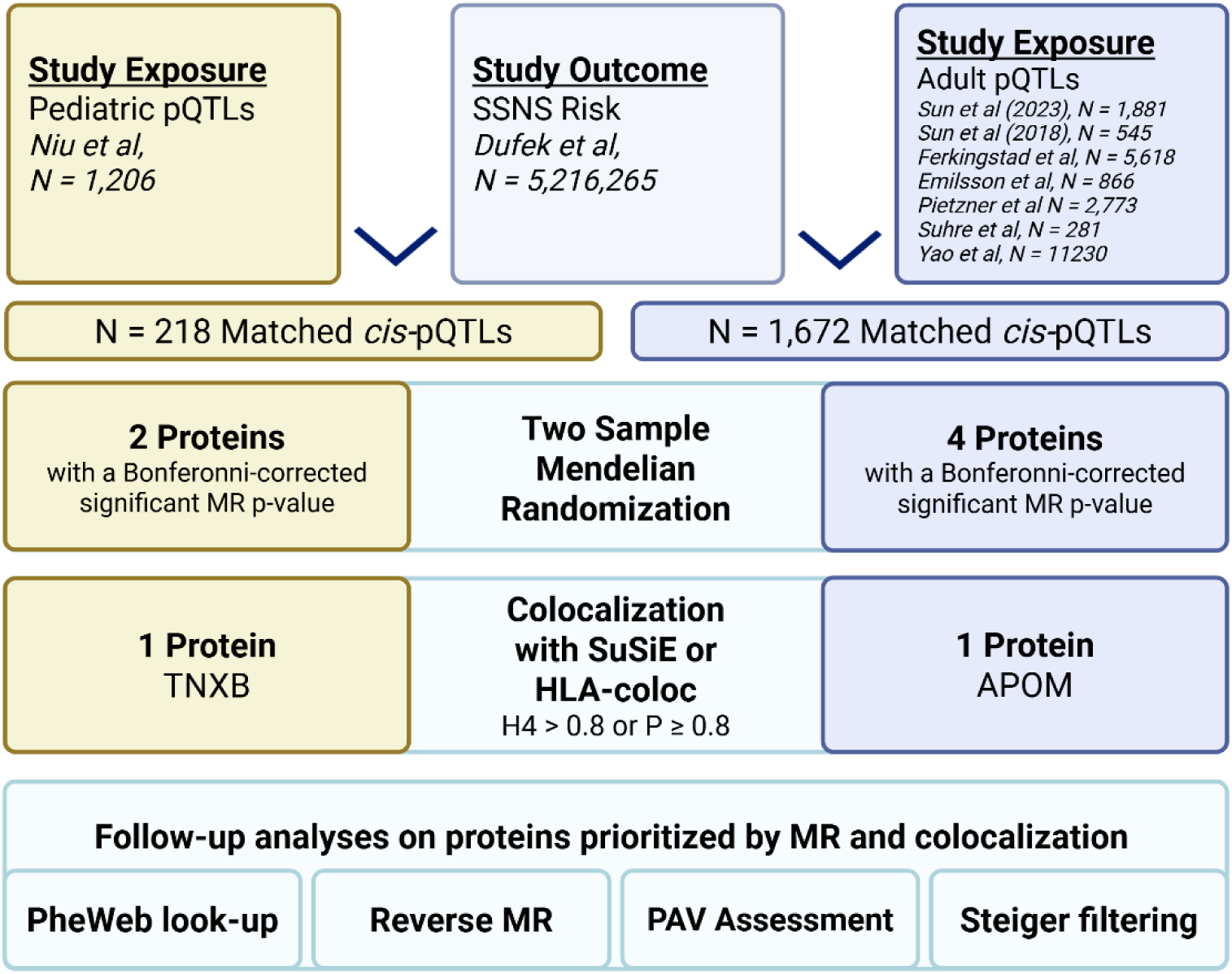
A schematic of the workflow and data sources used in the proteomic SSNS MR. Diagram includes breakdown of pediatric and adult pQTL studies, with the total number of statistically significant *cis-*pQTLs identified in each, as well as the SSNS GWAS and the number of reported SNVs. After filtering and selecting for genome-wide significant, independent, and valid pQTLs, 218 SNVs were matched with outcome data in children, and 1,672 SNVs were matched in adults. MR was conducted using the TwoSampleMR package, and colocalization was attempted with coloc and HLAcoloc packages.^10,35,37^ Further functional analysis was then done on proteins supported by MR and colocalization.

**Figure 2.**
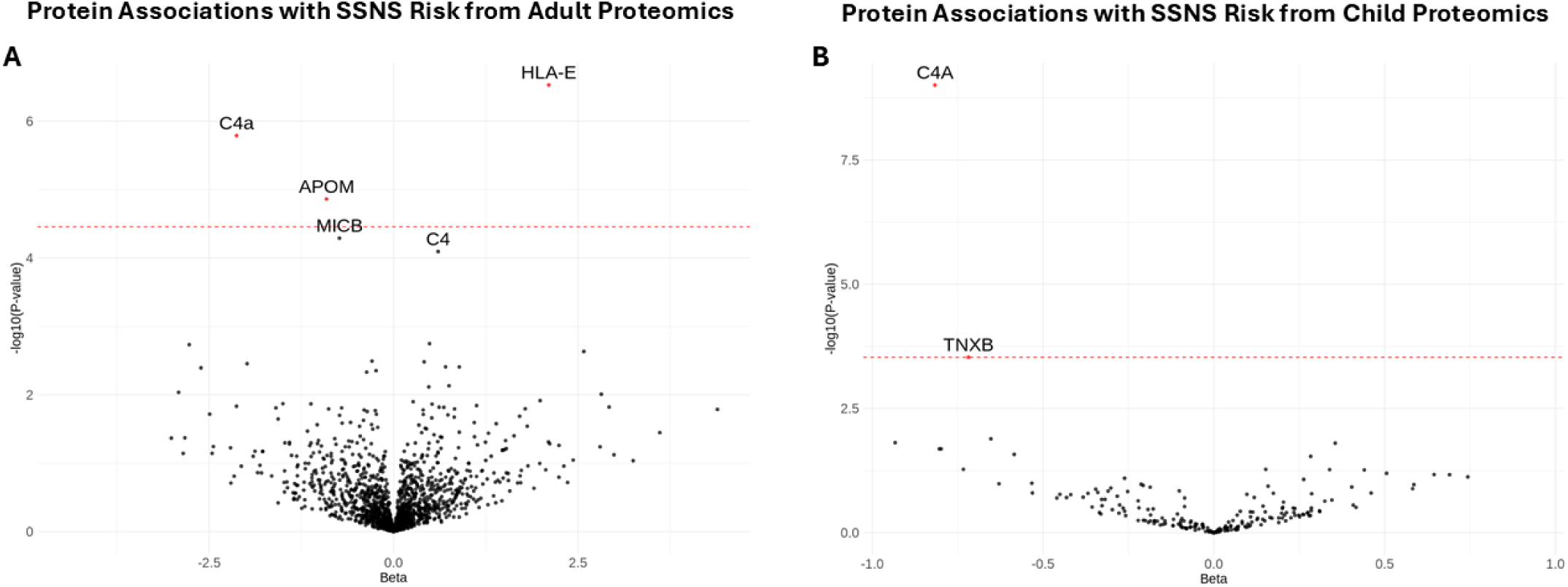
Volcano plot showing the association between protein levels and SSNS risk using (A) adult proteomic studies and (B) child proteomic studies. The x-axis represents effect size (beta), and the y-axis shows the –log10(p-value) of the association. The dotted red line marks the significance threshold based on Bonferroni correction for the number of unique proteins tested. Statistically significant (red points) and near-significant proteins are labelled. APOM and HLA-E were found to be associated with SSNS using adult proteomic studies, TNXB was associated with SSNS using child proteomics, and C4A was significantly associated with SSNS risk using all proteomic studies.

**Table 1.** Results of MR using the Wald ratio for pQTLs found to be significantly associated with SSNS in adults. 1,465 unique proteins were tested, resulting in a Bonferroni-adjusted significance threshold of *p* = 3.4 × 10^−5^. b: beta, se: standard error, p: p-value, OR: odds ratio, 95% CI: 95% confidence intervals. APOM was the only protein with a significant p-value that passed Steiger filtering.

| SNV | b | se | p | steiger_dir | steiger_pval | Protein | OR | 95% CI |
| --- | --- | --- | --- | --- | --- | --- | --- | --- |
| rs2853999 | 2.103 | 0.410 | 2.95E-07 | FALSE | 2.99E-58 | HLA-E | 8.190 | 3.67 - 18.30 |
| rs3117580 | -2.129 | 0.444 | 1.63E-06 | FALSE | 2.53E-18 | C4a | 0.119 | 0.05 - 0.28 |
| rs805270 | -0.909 | 0.209 | 1.37E-05 | TRUE | 1.89E-01 | APOM | 0.403 | 0.27 - 0.61 |

**Table 2.** Results of MR using the Wald ratio for pQTLs found to be significantly associated with SSNS in children. 171 unique proteins were tested, resulting in a Bonferroni-adjusted significance threshold of *p* = 2.9 × 10^−4^. b: beta, se: standard error, p: p-value, OR: odds ratio, 95% CI: 95% confidence intervals. TNXB was the only protein with a significant p-value that passed Steiger filtering.

| SNV | b | se | p | steiger_dir | steiger_pval | Protein | OR | 95% CI |
| --- | --- | --- | --- | --- | --- | --- | --- | --- |
| rs389884 | -0.817 | 0.134 | 9.82E-10 | FALSE | 5.73E-01 | C4A | 0.442 | 0.34 - 0.57 |
| rs9267671 | -0.719 | 0.198 | 2.95E-04 | TRUE | 3.69E-03 | TNXB | 0.487 | 0.33 - 0.72 |

To assess whether our MR-prioritized pQTLs were robust indicators of protein exposure, we examined whether the same SNVs were identified as pQTLs for the corresponding proteins across independent proteomic studies and measurement platforms.. The MR-prioritized *cis*-pQTLs for C4A were from the Soma-Logic based Pietzner et al study in adults and mass spectrometry-based Niu et al in children (rs3117580 and rs389884, respectively, R^2^ : 0.43). The *cis-*pQTL tested for APOM was identified by Sun et al (2023) on O-link, and a shared signal was detected in Ferkingstad et al using SomaLogic (rs805270 and rs805295, respectively, R^2^ : 0.89). The *cis-*pQTL tested for TNXB was identified by Niu et al with mass-spectrometry, and a related signal was detected in Sun et al (2023) on O-link (rs9267671 and rs9267797, respectively, R^2^ : 0.25). A *cis-* pQTL for HLA-E was not detected in any studies other than Sun et al (2023). Therefore, there was cross-study and cross-platform support for the precision and robustness of most of the MR-prioritized pQTLs as indicators of protein exposure.

We then applied Steiger filtering to the *cis*-pQTLs of the MR-prioritized proteins and found that APOM and TNXB had the only *cis*-pQTLs in which Steiger directionality was true (Supplementary Data 4 and 7). Thus, APOM and TNXB were the only proteins retained for subsequent analysis. Reverse MR was performed using SSNS as exposure and candidate proteins as outcomes. For these analyses, we used 16 genome-wide significant (p<5×10^−8^) independent (LD R^2^<0.01) SNVs as instruments for SSNS. Reverse MR analyses were restricted to APOM and TNXB, both of which had full summary levels GWAS from Sun et al (2023) and Niu et al, respectively.^16,22^ For each reverse MR, we performed multi-instrument two sample MR using the inverse variance weight method, as well as four pleiotropy-robust methods (MR-Egger, weighted median, weighted mode, MR-PRESSO). There was no evidence of reverse causation in either reverse MR with APOM and TNXB (Supplementary Data 8 and 9).

### PheWeb search

We searched the PheWeb database to look up GWAS associations of the *cis*-pQTLs of the MR-prioritized proteins for possible confounding factors. The *cis*-pQTL for TNXB (rs9267671) had a genome-wide significant association with psoriasis, and the APOM pQTL (rs805270) had no significant associations. The *cis*-pQTLs for TNXB and APOM both had suggestive associations (1×10^−5^<p<5×10^−8^) with celiac disease. Neither *cis*-pQTL had any associations with SSNS or other kidney disease phenotypes means less chance our MR result is affected by confounding.

### Colocalization

Both APOM and TNXB are located within the HLA region on chromosome 6. We first attempted to colocalize candidate proteins with SSNS using summary level SNV data. These results were unreliable and imprecise as colocalization would often fail to compile the data due to extensive LD in the region. Thus, we employed a new method of genetic colocalization at the HLA locus, using HLA alleles instead of SNVs.^37^

Using HLA colocalization, we found that APOM had a 97.8% probability of colocalization at *HLA-DRB1*, and TNXB had an 80.2% probability of colocalization at *HLA-DQA1* (Figure 3 and Supplementary Data 10-11). These results suggest that there is a shared signal at these respective HLA alleles that is causal for both protein level and SSNS risk.

**Figure 3.**
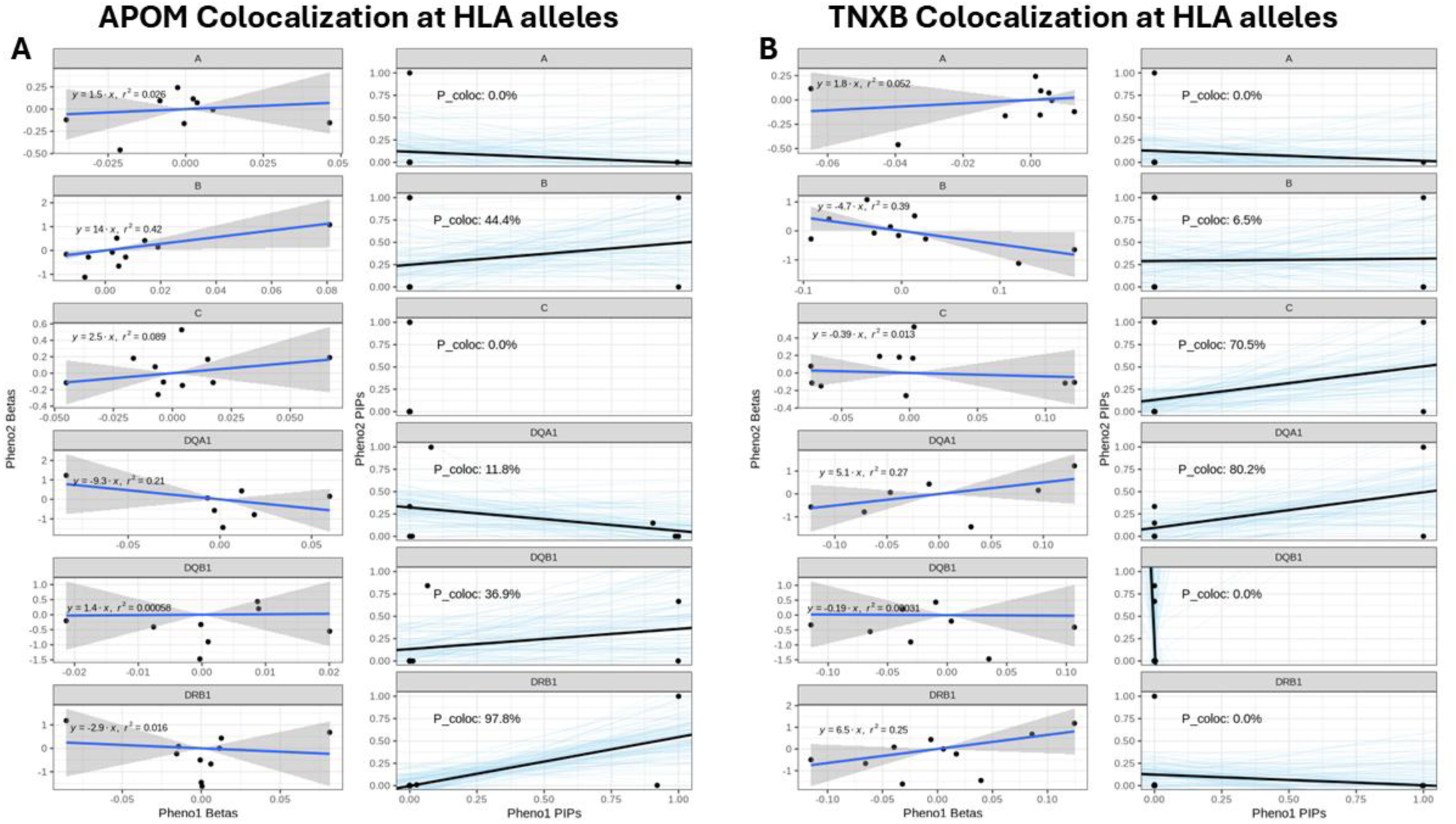
Results from HLAcoloc analysis for APOM (A) and TNXB (B). The left subfigures show the effect size of HLA alleles on the protein level in the blood (Pheno1) on the x-axis, and the relative effect on SSNS risk (Pheno2) on the y-axis. The right subfigures compare the posterior inclusion probabilities (PIPs) of HLA alleles for protein level on x-axis, and SSNS risk on the y-axis. The probability of colocalization of APOM at HLA-DRB1 is 97.8%, and the probability of colocalization on TNXB at HLA-DQA1 is 80.2%. This suggests that variation in HLA-DRB1 and HLA-DQA1 is causal for altered APOM/TNXB levels as well as increased SSNS risk.

Given the high degree of linkage disequilibrium patterns as HLA, we hypothesized that we may have obtained a false positive colocalization result if the *cis*-pQTL (SNV) used in our MR was in high LD with the HLA allele-pQTL for the same tested protein. Thus, as a quality check, we tested for LD between *cis*-pQTLs and HLA-pQTLs and found that they were in linkage equilibrium for both MR-prioritized proteins (Supplementary Table 12 and 13).

### PAV Assessment

*Cis-*pQTLs of APOM and TNXB were annotated with Ensembl VEP v115 to assess if they were protein altering variants (PAVs).^43^ PAVs were defined as coding sequence variants, frameshift variants, in-frame deletions, in-frame insertions, missense variants, protein altering variants, splice acceptor variants, splice donor variants, splice region variants, start lost, stop gained, or stop lost variants.^23^ Neither of the *cis*-pQTLs for APOM and TNXB (rs805270 and rs9267671) were found to be protein altering. We then investigated the variants in LD (R^2^ > 0.1) with the *cis*-pQTLs for a protein-altering effect. In APOM, rs707922 was found to be a missense variant (R^2^ with APOM pQTL: 0.51). rs707922 has not been found to be genome-wide significant for any disease phenotypes, and with a SIFT score of 0.2 and PolyPhen score of 0, it is unlikely to impair protein function. In TNXB, rs41258944 was also found to be a missense variant (R^2^ with TNXB pQTL: 0.51). rs41258944 has only been found to be genome-wide significant for psoriasis phenotypes and has a SIFT score of 0.17 and PolyPhen score of 0.11. However, the presence of both of these missense variants suggest that these *cis*-pQTLs could be subject to potential binding effects (Supplementary Table 14 and 15).

### eQTL Assessment

From the Genotype-Tissue Expression (GTEx) Portal,^33^ pQTLs for APOM and TNXB were both found to be eQTLs for several genes, however neither were eQTLs for APOM or TNXB. rs9267671 (TNXB *cis*-pQTL) was found to alter expression levels of *ZBTB12* in whole blood, *NOTCH4* in adipose tissue, and *MICB* in multiple tissues and whole blood. The TNXB *cis*-pQTL is also an eQTL of *TNXA*, a pseudogene of TNXB, but this effect was only detected in the adrenal gland and tibial nerve. rs805270 (APOM *cis*-pQTL) is an eQTL of the neighbouring gene *LY6G5B* in many different tissues and in whole blood, as well as a blood eQTL of *AIF1*.

### Kidney Cell Atlas

We queried the Kidney Cell Atlas to better understand which kidney cell types highly express TNXB and APOM. APOM was found to be highly expressed in proximal tubular cells, and TNXB was highly expressed in peritubular capillary endothelial and vasa recta endothelial cells.

### HLA allele binding assessment

For each HLA allele identified by colocalization for MR-prioritized proteins, we evaluated binding affinity of peptides (eluted ligands score) within these proteins using the NetMHCIIpan resource.^40^ For APOM, which colocalized at *HLA-DRB1*, NetMHCIIpan showed that *HLA-DRB1* had a greater total amount of interactions (including strong and weak) for APOM peptides compared to other class II HLA alleles (Supplementary Data 16). TNXB, which colocalized at *HLA-DQA1*, was also found to have a greater total amount of interactions (strong and weak) with *HLA-DQA1* than other class II genes (Supplementary Data 17). NetMHCIIpan supported the finding that *HLA-DRB1* and *HLA-DQA1* could be interacting with the peptides of TNXB and APOM to disrupt protein levels.

### Power calculation

Power of our MR study was estimated at 80% to detect an Odds Ratio of 2.2 (of the protein being associated with SSNS), since the proportion of cases (children with SSNS) was 7% in the outcome SSNS GWAS of 6,064 individuals,^25^ and the median variance explained per protein in Europeans is 4.6%.^23^ Power was computed using an alpha = 0.05/1,465 = 3.4×10^−5^ (Bonferroni correction).^44^

Our findings by two sample MR and HLA colocalization suggest that reduced blood TNXB and APOM levels are causally associated with SSNS, indicating TNXB and APOM as potential targets or biomarkers for SSNS risk.

## DISCUSSION

In this study, we evaluated the causal effects of 1,540 proteins on SSNS risk and found that lowered APOM and TNXB levels are causally associated with SSNS susceptibility in Europeans using two-sample MR. Colocalization analysis found that TNXB and APOM have shared genetic signal with SSNS at *HLA-DQA1* and *HLA-DRB1*, respectively, and that peptides from these proteins have higher binding affinity to their respective HLA alleles. Genes annotated to the *cis*-pQTLs for APOM and TNXB are widely expressed through the body. These results suggest that dysregulation of TNXB or APOM levels in the blood could modify the risk for SSNS, and that these proteins could serve as actionable targets for treatment of disease, or biomarkers for risk prediction.

### Apolipoprotein M (APOM)

Our analyses prioritized APOM as a functionally relevant protein in SSNS. APOM is an apolipoprotein which comprises a family of molecules that have a variety of functions including providing structural stability and assisting transport of lipoproteins, activating inflammatory pathways, and acting as chaperones for vitamins and lipids, among others.^45,46^ The plasma concentration of APOM has previously been implicated in a variety of kidney diseases via observational studies, including nephrotic syndrome.^47^ Plasma APOM levels were found to be lower in patients with idiopathic nephrotic syndrome and end stage renal disease compared to healthy controls,^48–50^ and were found to negatively correlate with chronic kidney disease severity.^50^ In nephrotic syndrome in particular, APOM levels positively correlated with albuminuria,^48^ but no difference in APOM levels were observed when comparing patients with variety degrees of albuminuria secondary to diabetic nephropathy.^51^ There is also growing evidence to support the causal relationship between APOM and kidney disease, with reduced APOM levels resulting in increased kidney injury.^52^ Similarly, overexpression of APOM in mice with IgA nephropathy (HIGA mice) resulted in less kidney injury and lower expression of markers of fibrosis compared to healthy animals.^53^

Importantly, APOM is the principal chaperone of a bioactive lipid called sphingosine-1-phosphate (S1P),^54^ which acts by binding to 5 different receptors, 4 of which are expressed in podocytes.^47^ Studies using mouse models have found that disruption or knockouts in the APOM-S1P pathway leads to proteinuria and podocyte damage.^55–57^ In humans, sphingosine-1-phosphate lyase (SGPL1) deficiency, which results in increased S1P levels in the blood, has been associated with steroid resistant nephrotic syndrome.^58,59^ The mechanisms by which S1P bound to APOM lead to proteinuria are unclear. Indeed, perhaps S1P acts directly on podocytes. Alternatively, S1P plays a role in the immune system, which may then target the kidney. S1P has been shown to trigger migration of T cells from the thymus into the bloodstream.^60^ This T-cell migration is dependent on the gradient of S1P between lymphoid organs (such as the thymus and lymph nodes) and the lymphatic and blood vessels, and in this way can regulate immune response.^60^ If S1P signalling becomes imbalanced, T-cell response can be altered. For example, in lupus nephritis, another proteinuric renal disease, S1P signaling can become imbalanced, leading to increased number of pathogenic T cells migrating to the kidneys.^61^ As a small lipid molecule, we could not test if S1P abundance levels were causal in SSNS as there are no pQTLs for S1P.

To date, there are multiple approved drugs targeting S1P receptors, such as Fingolimod and Ozanimod. Both of these drugs act as functional antagonists, by binding and activating a variety of S1P receptors.^62,63^ Fingolimod targets S1P receptors 1, 3, 4, and 5,^63^ whereas Ozanimod selectively targets receptors 1 and 5.^64^ Receptors 1 through 4 are expressed in podocytes and all are expressed in immune cells, including lymphocytes.^47,65^ These drugs are currently approved to treat other autoimmune disorders, such as multiple sclerosis, and have shown promising results in treating psoriasis.^66,67^ Fingolimod modulates sphingosine-1-phosphate receptors on lymphocytes, preventing their entry into the central nervous system and other tissues, thereby dampening the immune response.^68^ In patients with multiple sclerosis (MS), a weaker immune response reduces the inflammation and nerve damage caused by immune cells attacking the protective myelin around neurons.^68^ Perhaps a similar effect could be achieved for children with nephrotic syndrome. This evidence supports the potential repurposing of these drugs for the treatment of SSNS by targeting S1P receptors and reducing immune responses that may be directed against the kidney.

### Tenascin XB (TNXB)

TNXB is part of a family of large extracellular matrix proteins.^69^ It is a glycoprotein which is expressed in connective tissues, including the kidney glomeruli.^70^ There is some evidence that mutations in *TNXB* can lead to kidney phenotypes, but these reflect rare variants causing congenital anomalies of the kidney and urinary tract (CAKUT) or systemic conditions such as Ehlers-Danlos syndrome.^71,72^ To date, there has been no evidence of variants within *TNXB* or expression levels of the TNXB protein having an effect on SSNS or proteinuria phenotypes. There are no currently approved drugs targeting TNXB or its related pathways.

### Colocalization of proteins and SSNS via *HLA-DRB1* and *HLA-DQA1*

Colocalization using HLA alleles rather than SNVs has allowed us to interrogate the highly polymorphic and complexly linked HLA locus, which is the genetic locus most strongly associated with SSNS. This analysis has allowed us to translate the genetic findings within this region into candidate therapeutic targets. We found shared signal at the two most strongly associated HLA alleles from previous GWAS in SSNS: *HLA-DRB1* and *-DQA1.*^25,73–77^ This implies that these HLA alleles are likely causal for SSNS. We subsequently evaluated binding affinity of peptides comprising our MR-prioritized proteins to their respective HLA alleles and found increased binding affinity of APOM with *HLA-DRB1* and TNXB with *HLA-DQA1.* Our results support the prioritization of -*DRB1* and -*DQA1* presented peptides as drug targets for modulating SSNS development

### Child- versus adult-derived proteomics studies

We found different MR-prioritized proteins between the child-derived proteomics GWAS and the adult-derived proteomics GWAS as exposures. Indeed, longitudinal studies report a differing proteomic profile across the lifespan,^78^ and diverse factors including age, sex, growth, body mass index, and puberty, can affect the levels of circulating proteins.^79^ As the typical age of onset of SSNS is between 2-5 years of age,^1^ examining circulating proteins by age may be relevant. SSNS is often interchangeable with the adult-onset minimal change disease, and thus the proteins identified from the adult-derived proteomics data may be more relevant in older children/young adults. In contrast, the study by Niu et al found that 92% of the *cis-*pQTLs identified in children were successfully replicated in adults.^22^ Further large-scale proteomic studies in children are warranted to better characterize the protein landscape across the lifespan. Lastly, as a point of strength, the APOM pQTL was replicated in multiple studies we analyzed, with either the same SNV or a SNV in high LD significantly associated with altered levels of the same protein (LD-linked pQTLs of APOM were found in Sun et al [2023], Emilsson et al, Ferkingstad et al, Pietzner et al, and Sun et al [2018]). However, other pQTLs of TNXB were in weak or low LD with the MR-prioritized pQTL, making it unclear if they represent the same genetic signal or not. It is possible that the effects of genetic variation on TNXB abundance vary across the lifespan.

### Limitations

Our study had limitations. First, our study tested the effect of circulating plasma proteins and did not account for potential tissue-specific effects. Second, we did not validate our candidate MR proteins in an independent cohort of SSNS cases versus controls, since this was beyond the scope of the current study. Third, the HLA-pQTLs used for colocalization were obtained from an independent HLA-specific proteomics study rather than from the eight SNV-based *cis*-pQTL studies used to conduct the MR analyses. This meant that some MR-prioritized proteins were not examined by colocalization, since they did not have a genome-wide significant HLA-pQTL (e.g. C4A and HLA-E). Finally, our MR and colocalization studies were performed in European populations, which limits the generalizability of our findings to populations of other ancestries. This underlines the importance of further recruitment and analysis of non-European patients in large-scale biobanks going forward.

## CONCLUSION

In conclusion, our MR study found that genetically altered levels of APOM and TNXB are causally associated with SSNS development in European individuals. These proteins were further validated by colocalization analysis identifying causally associated HLA alleles at *HLA-DRB1* and *HLA-DQA1*. APOM and TNXB are promising biomarkers for early identification of the SSNS and are also compelling new drug targets for the treatment of disease. Repurposing of currently available drugs targeting the APOM/S1P pathway could be considered. Future studies are needed to validate these proteins as biomarkers in an independent case-control SSNS cohort.

## Data Availability

All data produced in the present study are contained in the supplementary data or available upon request to the authors.

https://pubmed.ncbi.nlm.nih.gov/37794186/

https://pubmed.ncbi.nlm.nih.gov/34857953/

https://pubmed.ncbi.nlm.nih.gov/30072576/

https://pubmed.ncbi.nlm.nih.gov/34648354/

https://pubmed.ncbi.nlm.nih.gov/28240269/

https://pubmed.ncbi.nlm.nih.gov/30111768/

https://pubmed.ncbi.nlm.nih.gov/39972214/

https://pubmed.ncbi.nlm.nih.gov/29875488/

https://pubmed.ncbi.nlm.nih.gov/31263063/

https://pubmed.ncbi.nlm.nih.gov/39085222/

## ACKNOWLEDGEMENTS

We would like to acknowledge the patients and families that contributed to this research without direct benefit to themselves. We would also like to thank Professors Horia Stanescu, Detlef Bockenhauer, Daniel Gale, and Robert Kleta for their mentorship and guidance in this project.

M.L. Downie is supported by the KRESCENT program from the Kidney Foundation of Canada and the Canadian Institute of Health Research Early Career Researcher operating grant for this work.

